# Pretreatment and interventional parameters predict excellent recanalization of large vessel occlusion in patients with acute ischemic stroke

**DOI:** 10.1101/2023.08.22.23294452

**Authors:** Vivek Yedavalli, Manisha Koneru, Meisam Hoseinyazdi, Cynthia Greene, Karen Copeland, Risheng Xu, Licia Luna, Justin Caplan, Adam Dmytriw, Adrien Guenego, Jeremy Heit, Gregory Albers, Max Wintermark, Fernando Gonzalez, Victor Urrutia, Judy Huang, Richard Leigh, Elisabeth Marsh, Rafael Llinas, Argye Hillis, Kambiz Nael

## Abstract

**Background:** In patients with acute ischemic stroke secondary to large vessel occlusion (AIS-LVO), improved functional outcomes have been reported in patients who achieve Modified Thrombolysis In Cerebral Infarction (mTICI) 2c/3 (excellent recanalization) over mTICI 2b. We aimed to determine pretreatment and interventional variables that could predict achieving mTICI 2c/3 over 2b reperfusion in patients who underwent technically successful mechanical thrombectomy (MT).

**Methods:** In this retrospective study, consecutive AIS patients with anterior circulation LVO who underwent MT and achieved recanalization with mTICI 2b/2c/3 were included. We evaluated the association between pretreatment clinical and imaging variables and interventional parameters in patients who achieved mTICI 2c/3 vs. 2b using logistic regression and ROC analyses.

**Results:** From 5/11/2019 to 10/09/2022, 149 consecutive patients met our inclusion criteria (median 70 years old [IQR 65 - 78.5], 57.7% female). Adjusted multivariate regression analyses showed that patients with excellent recanalization had lower admission NIHSS (aOR 0.93, p = 0.036), were less likely to have a history of diabetes mellitus (DM) (aOR 0.42, p = 0.050) and prior stroke (aOR 0.27, p = 0.007), had a cerebral blood volume (CBV) index >= 0.7 (aOR 3.75, p = 0.007), and were more likely to achieve excellent recanalization with aspiration alone (aOR 2.89, p = 0.012). A multivariate logistic regression model comprising these independent factors predicted mTICI 2c/3 with an AUC 0.79 (95% CI: 0.68-0.86; p < 0.001), sensitivity of 94%; specificity of 41%.

**Conclusion:** Robust collateral status (CS) defined by CBV index >= 0.7, absence of DM and prior stroke, lower initial stroke severity, and direct aspiration are all predictive of excellent recanalization in successfully recanalized AIS-LVO patients. Our findings highlight the prognostic implications of robust CS, DM and stroke prevention, as well as use of aspiration alone in maximizing the likelihood of excellent recanalization.

## Introduction

In patients presenting with acute ischemic stroke (AIS) due to anterior circulation large vessel occlusion (LVO), mechanical thrombectomy (MT) is now the standard of care for up to 24 hours after symptom onset.^1^ Successful recanalization, defined as modified thrombolysis in cerebral infarction (mTICI) scores of 2b or higher, is also associated with improved outcomes irrespective of the number of passes.^2^ Furthermore, the degree of recanalization even amongst the successfully recanalized patients plays a role in achieving functional independence. Prior studies have demonstrated that patients who achieve mTICI 2c and 3, also referred to as excellent recanalization, have early neurological improvement^3, 4^ and better clinical outcomes than those who achieve mTICI 2b.^3, 5–11^

However, few studies have directly investigated which pretreatment and interventional parameters are predictive of mTICI 2c/3 versus 2b in the successfully recanalized AIS-LVO group. Moreover, pretreatment imaging parameters are yet to be assessed for potentially predicting mTICI 2c/3 recanalization. Therefore, the aim of our study is to determine which pretreatment imaging, clinical, and interventional parameters may be predictive of mTICI 2c/3 recanalization. We hypothesize that more robust collateral status (CS) is associated with increased likelihood of achieving excellent recanalization.

## Materials and Methods

This study was approved by an institutional review board with waiver of informed consent. The corresponding author has full access to all data in the study and assumes responsibility for integrity and analyses. Data will be made available upon reasonable request to the corresponding author. This study was reported in accordance with Strengthening the Reporting of Observational Studies in Epidemiology (STROBE) guidelines.

### Study Population

We performed a retrospective cohort study of all patients presenting with AIS caused by a large vessel occlusion from 4/3/2019 to 5/25/2022 comprised of data from two comprehensive stroke centers. This study was approved by our institutional review board and complies with the Health Insurance Portability and Accountability Act.

Inclusion criteria were a) patients with AIS secondary to anterior circulation LVO (defined as distal intracranial ICA, M1, and proximal M2 segments of the middle cerebral artery [MCA]) who were evaluated within 24 hours of symptom onset, b) underwent CTP with postprocessed maps available on the Rapid software platform (iSchemaview, Menlo Park, CA), and c) successfully treated with MT to achieve mTICI 2b, 2c, or 3 recanalization. MT was performed by one of four experienced neurointerventionalists using FDA-approved thrombectomy devices at their discretion and in accordance with current technical standards.^12^ The mTICI was also determined by the performing neuro-interventionalist. Study participants were then categorized into either a mTICI 2b or a combined mTICI 2c/3 cohort for comparative analysis.

### Data Collection

Baseline and clinical data collected for each patient included demographics, risk factors for AIS including heart disease, hypertension, hyperlipidemia, diabetes mellitus (DM), atrial fibrillation, prior stroke, smoking status, alcohol status, history of malignancy, and body mass index (BMI), admission NIH stroke scale (admission NIHSS), anticoagulation status, site of occlusion, and laterality of occlusion. Heart disease was defined as inclusive of coronary artery disease (CAD), cardiomyopathy, congestive heart failure, and valvular disease.

Baseline lab parameters and vital signs were also collected including hemoglobin (Hb), glucose, creatinine clearance, blood urea nitrogen-to-creatinine ratio, systolic blood pressure (SBP), and diastolic blood pressure (DBP).

Additional collected parameters include IV thrombolytic administration, when applicable; door to needle time; door to groin puncture time; time from groin puncture to recanalization, door to recanalization time, number of passes, MT method (i.e., aspiration alone, stent retriever alone, or combined), and mTICI score.

The ASPECTS score, clot burden score (CBS), single phase CTA score based on Tan criteria13, multiphase CTA score on source CTP images (mCTA) based on the Menon mCTA collateral score^14^, and digital subtraction angiography (DSA) collateral score based on ASITN/SIR criteria^15^ were assessed by a board certified neuroradiologist (V.S.Y., 6 years of experience).

### Imaging Analysis

Whole brain CTP was performed with the following parameters: 70 kVP, 200 Effective mAs, Rotation Time 0.25 s, Average Acquisition Time 60 s, Collimation 48 x 1.2 mm, Pitch Value 0.7, 4D Range 114 mm x 1.5 seconds. CTP images are then post-processed using RAPID commercial software (IschemaView, Menlo Park, CA) for generating quantitative (relative cerebral blood flow (rCBF) < 20%, 30%, 34%, and 38%) and time to maximum (Tmax) volumes > 4 seconds, 6 seconds, 8 seconds, and 10 seconds. The hypoperfusion intensity ratio (HIR), defined as the ratio of the Tmax > 10 seconds and Tmax > 6 seconds volumes, was collected in addition to the cerebral blood volume index (CBV Index), which is defined as the relative CBV within the Tmax > 6 seconds volume.

Both centers within this study used the same CTP parameters.

### Study Outcomes

The primary outcome measure was achieving mTICI 2c/3 recanalization (excellent recanalization).

### Statistical Analysis

Patients were grouped by recanalization score (i.e., mTICI 2c/3 and mTICI 2b).

Continuous variables were reported as medians with interquartile range (IQR); categorical variables were reported as frequencies. Two-sided Student’s t-test, nonparametric Wilcoxon rank sum tests, chi-square test, or Fisher’s exact test were performed as applicable for between-group comparisons. Univariate regressions for outcome of mTICI 2c/3 vs. 2b were performed to yield unadjusted odds ratios (OR). Optimal cutoff point for CBV index was determined using receiver operating characteristics (ROC) curve and maximizing Youden’s J statistical index. Only significant univariate predictors were inputted into a multivariate forward stepwise logistic regression with greatest minimization of corrected Akaike information criterion (AICc) to derive adjusted odds ratios (aOR) for predictors that remained in the final model. Multivariate model parameters reported include AICc, Bayesian information criterion (BIC), and whole model chi-square test. Receiver operating characteristics (ROC) curve was bootstrapped with 2500 samples to yield area under the curve (AUC), sensitivity, and specificity with 95% confidence intervals (CI). Significance was pre-specified at 0.05.

## Results

A total of 419 consecutive patients with a vessel occlusion were identified in our database from 4/3/2019 to 5/25/2022. One hundred seventy-two were excluded because LVO was not present, MT was not attempted, or MT was unsuccessful. Ninety-eight were excluded because CTP information was not available at the time of imaging or imaging was nondiagnostic. A total of 149 patients (median, IQR 70 years old [64 - 78.5], 57.7% female) were included in this study.

Univariate regression analyses:

### Baseline characteristics

The location of LVO was internal cerebral artery (n=6, 4.0%); MCA-M1 (n=112, 75.2%); and MCA-proximal M2 (n=31, 20.8%). Patients in the excellent recanalization group had a lower prevalence of DM (22.7% versus 41.0%, p = 0.028) and prior stroke (16.4% versus 38.5%, p = 0.004), and they and presented with lower average admission NIHSS (median,15 [IQR 1 -19] versus 18 [IQR 13-23], p = 0.031). See Table 1.

**1.**
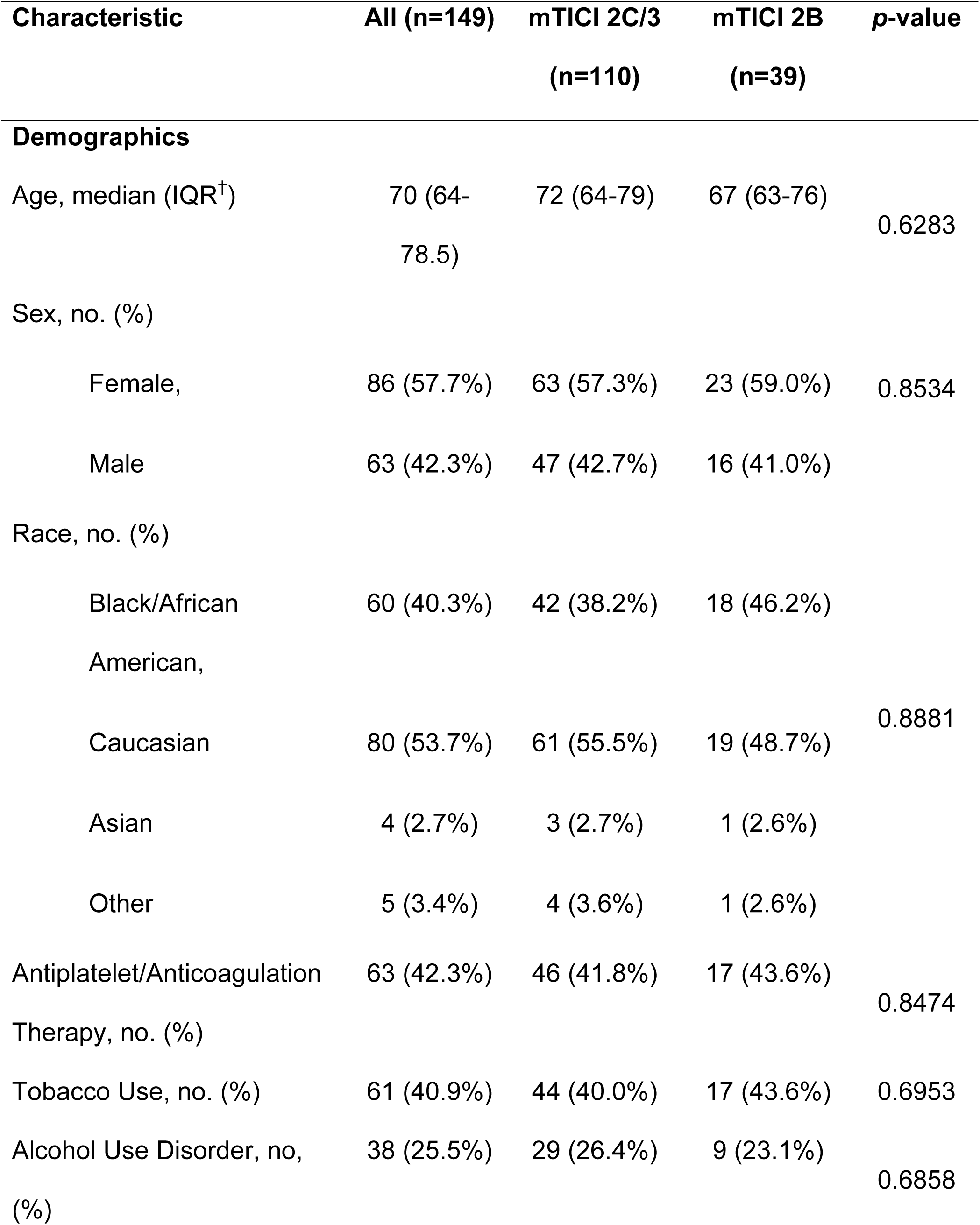
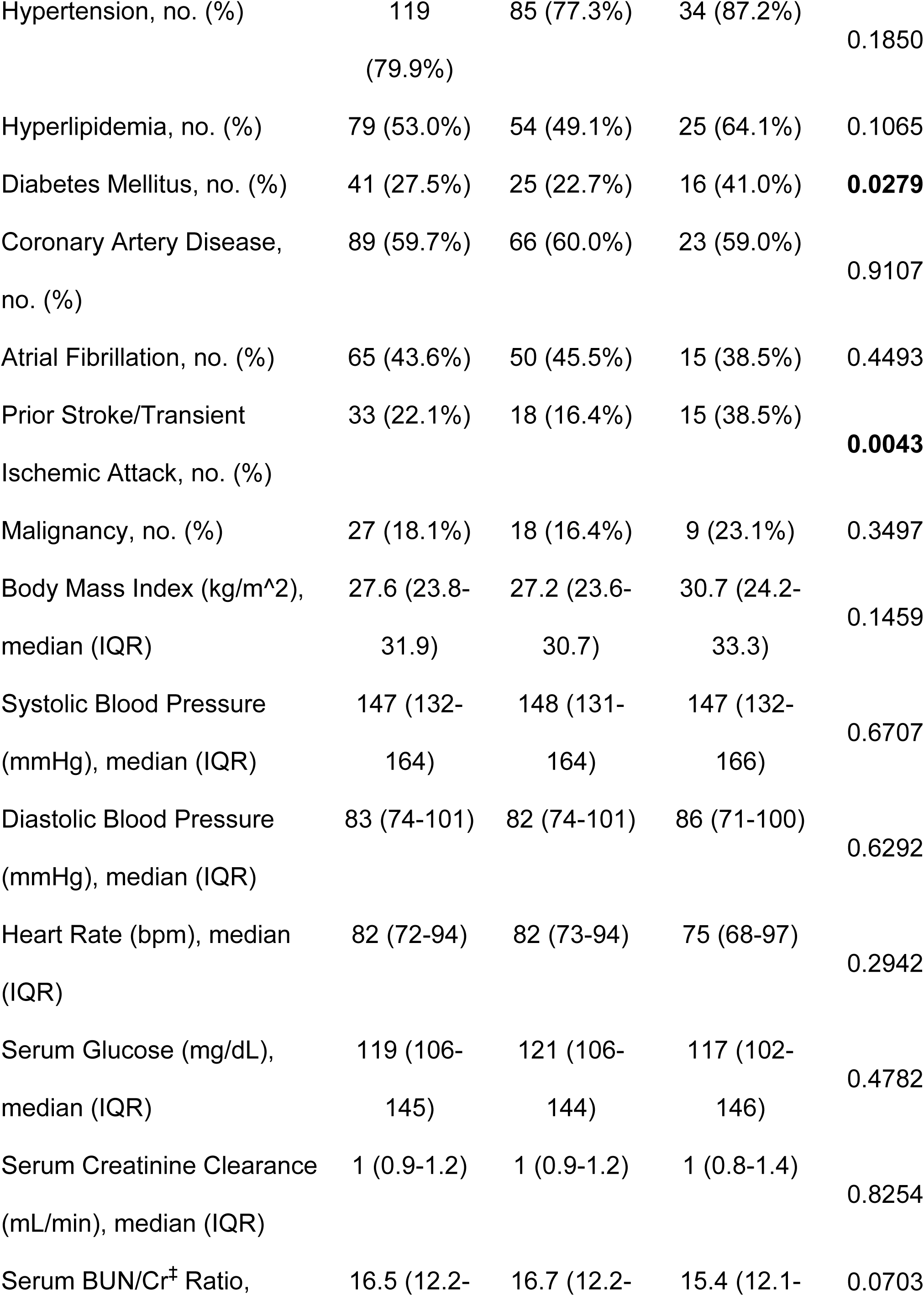
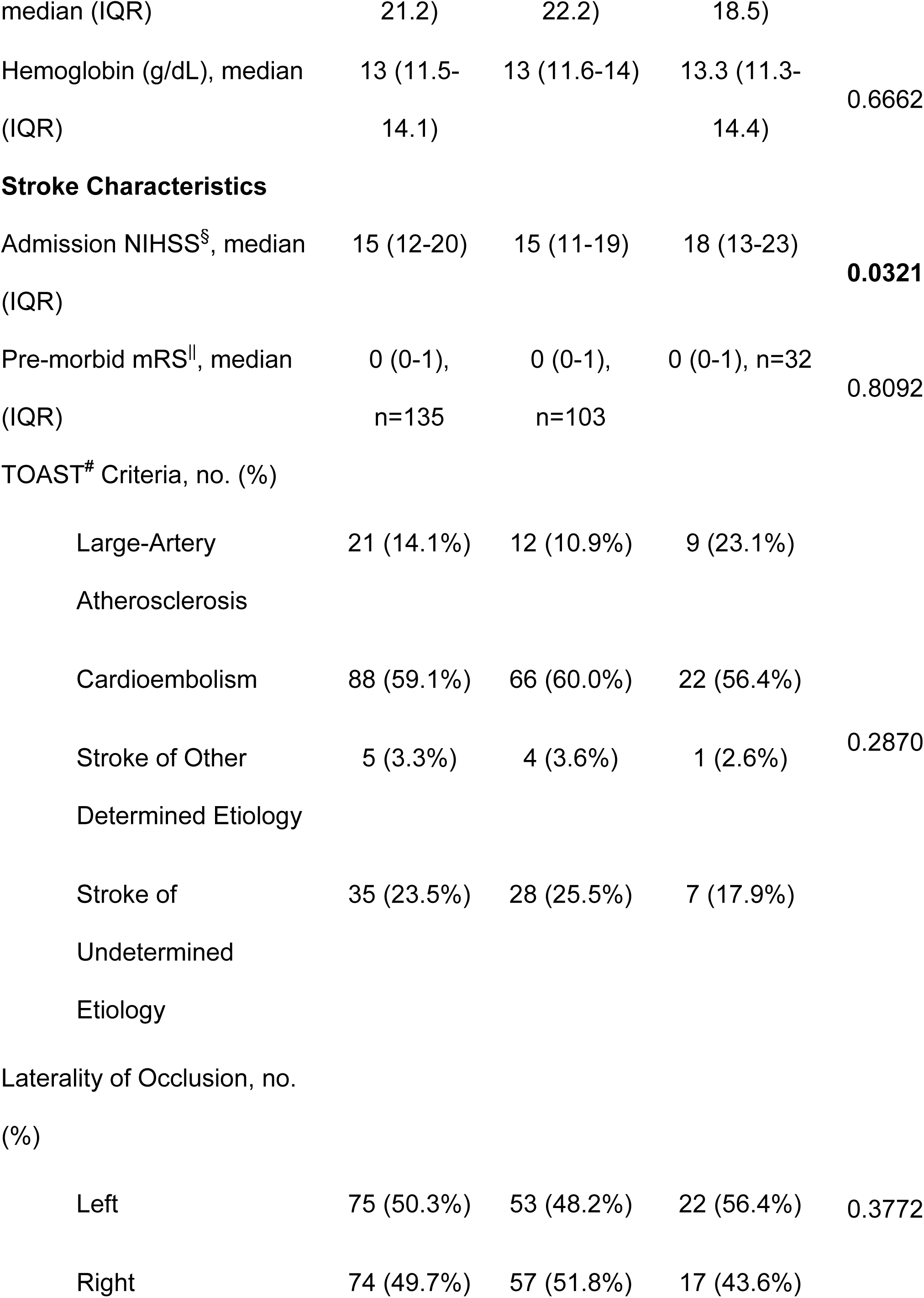
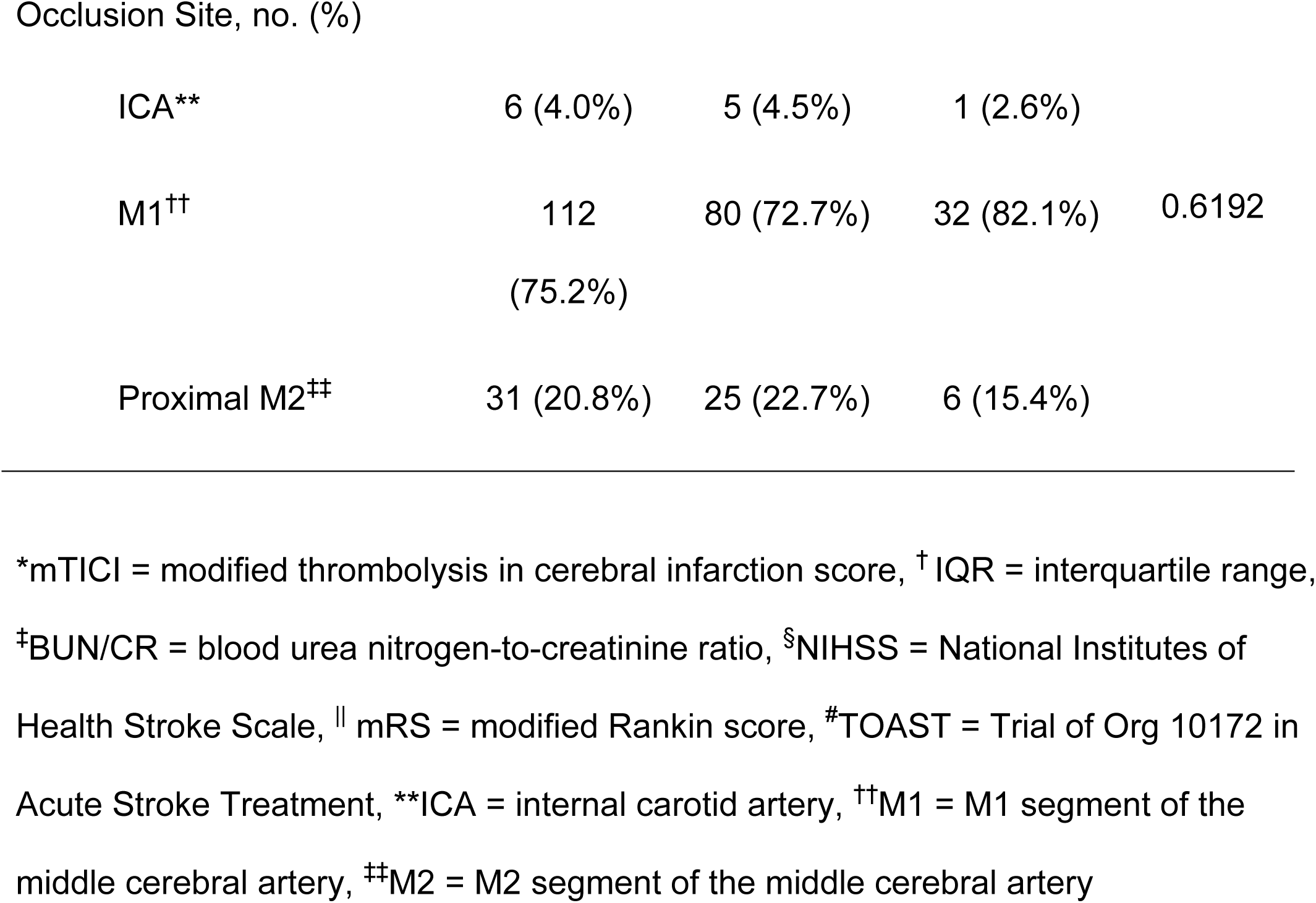
Baseline comparison between patients recanalized to mTICI* 2B vs. 2C/3.

### Imaging parameters

The relative cerebral blood flow (rCBF) volumes were lower in patients in the excellent recanalization group. The breakdown of rCBF volumes (median, IQR, ml) in patients with excellent recanalization vs. 2B were as follow: rCBF < 20% (0 ml, [0-5.3] versus 4 ml [0-31]; p = 0.001); rCBF < 30% (0 ml [0-17.3] versus 17 ml [0-67], p = 0.002); rCBF < 34% (7 ml [0-26] versus 22 ml [0-76], p = 0.005); rCBF < 38% (11 ml [0-34.3] versus 29 ml [5-87], p = 0.005). CBV index with a threshold of >= 0.7 was higher in the mTICI 2c/3 group (97/110, 88.2% versus 25/39, 64%, p < 0.001). See Table 2.

**2.**
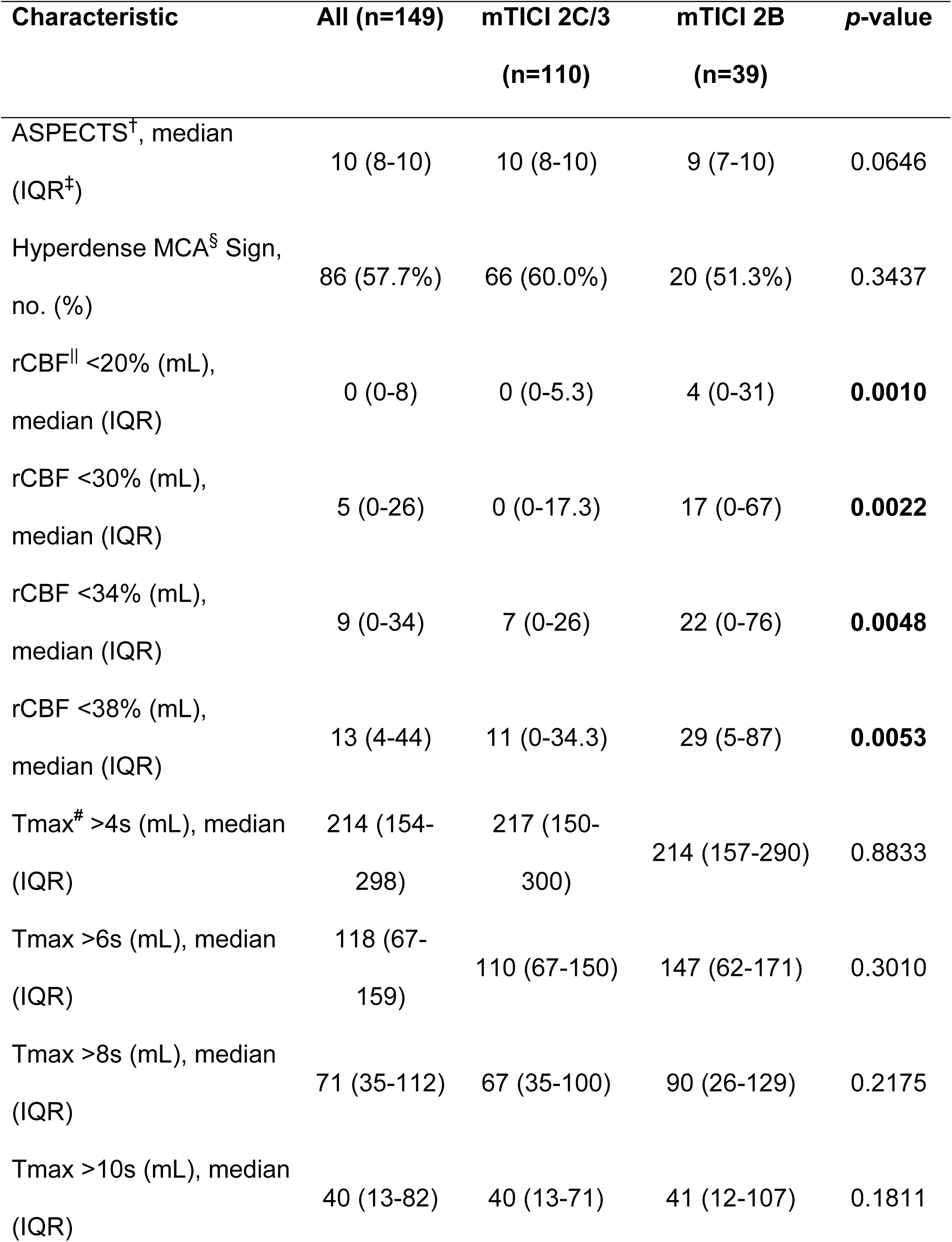
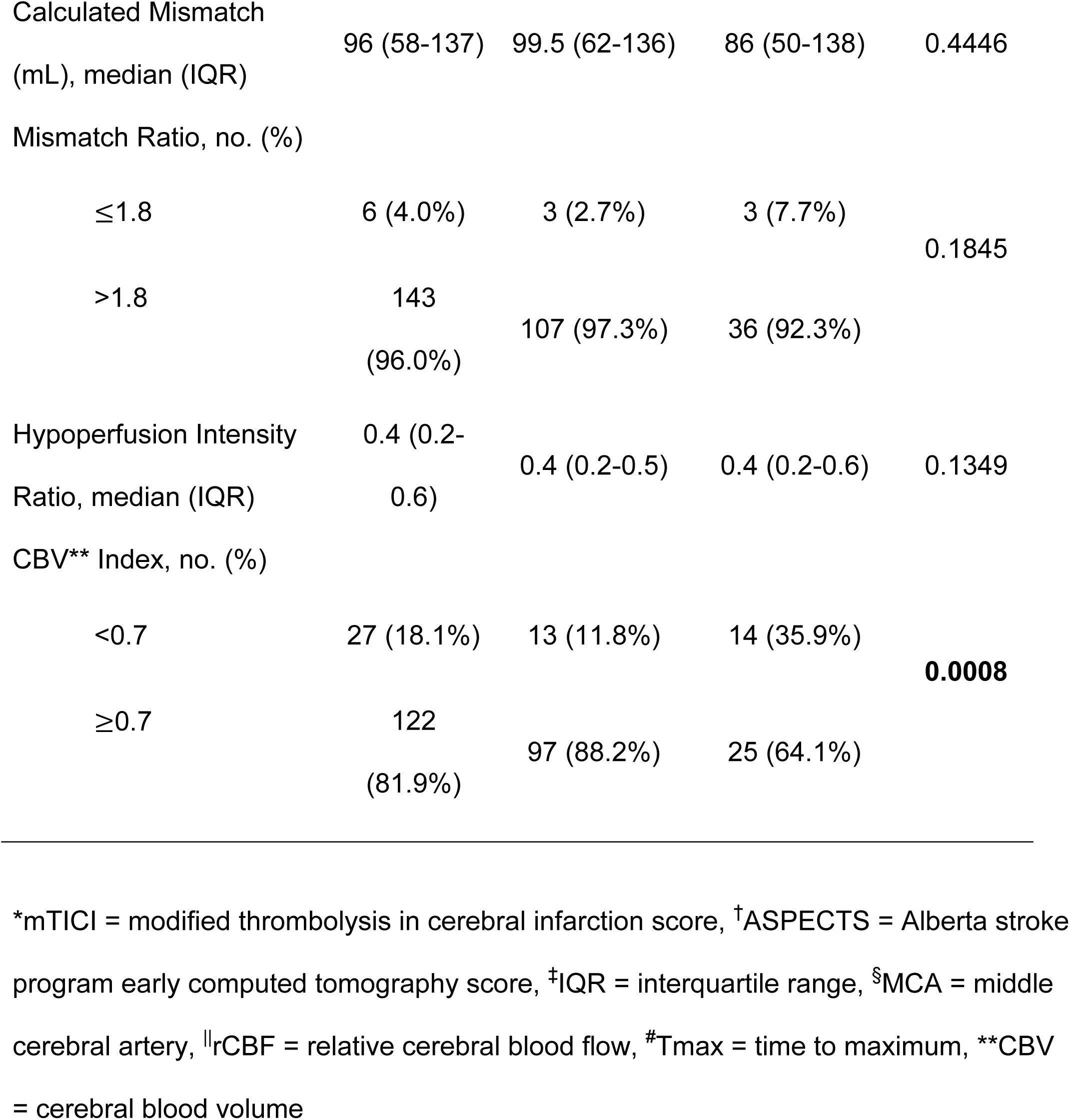
Imaging parameters between patients recanalized to mTICI* 2B vs. 2C/3.

### Interventional parameters

Patients with excellent recanalization had fewer passes (median 1, [IQR 1-1] versus 1 [IQR 1-2], p = 0.015), higher rate of direct aspiration MT (65.4% versus 35.9%, p = 0.005), lower rates of combined aspiration and stent retriever (29.1% versus 56.4%, p = 0.005) and shorter groin puncture to recanalization time (minutes) (median 32 minutes, [IQR 22-47] versus 48 [IQR 26-78], p = 0.002). See Table 3.

**3.**
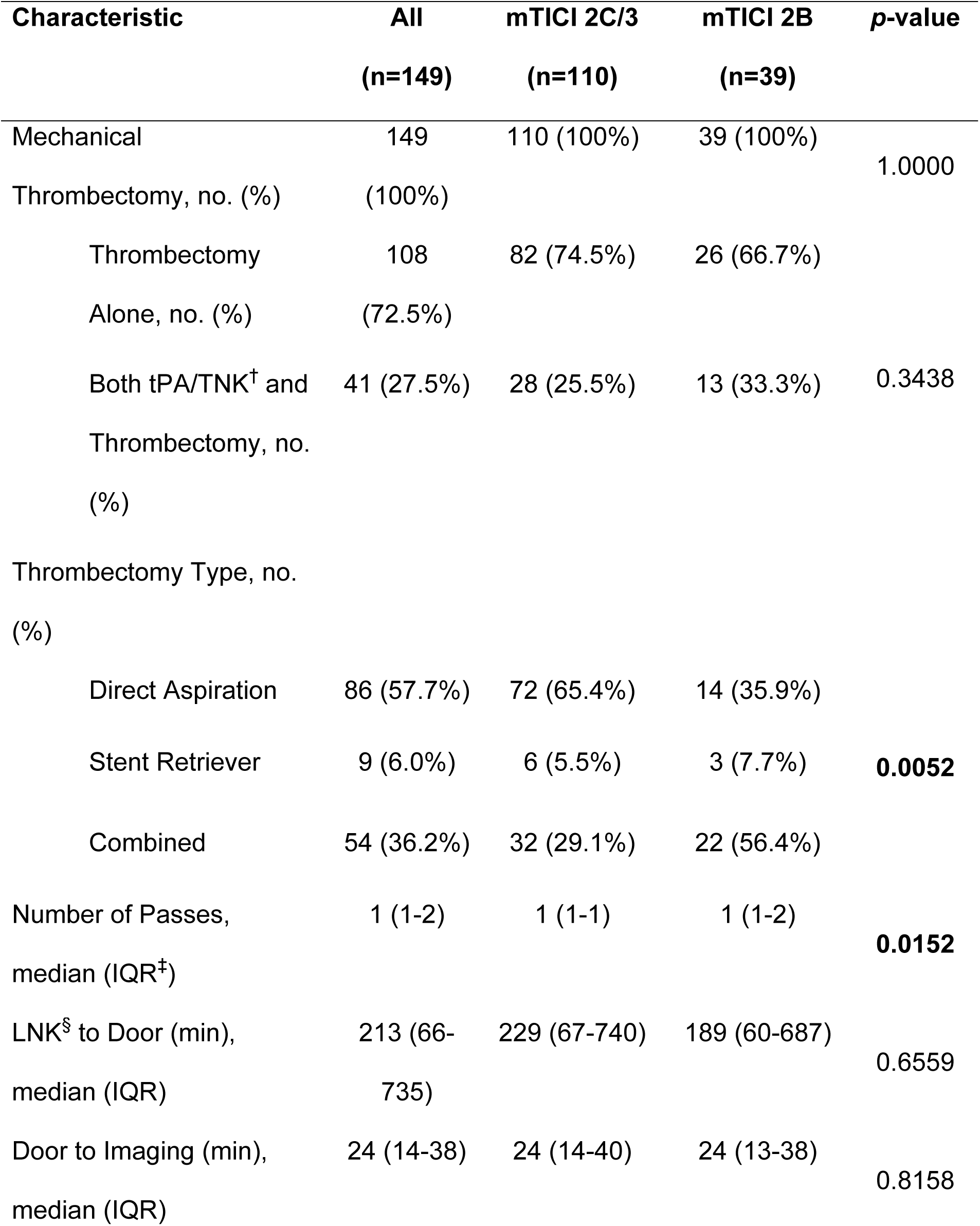
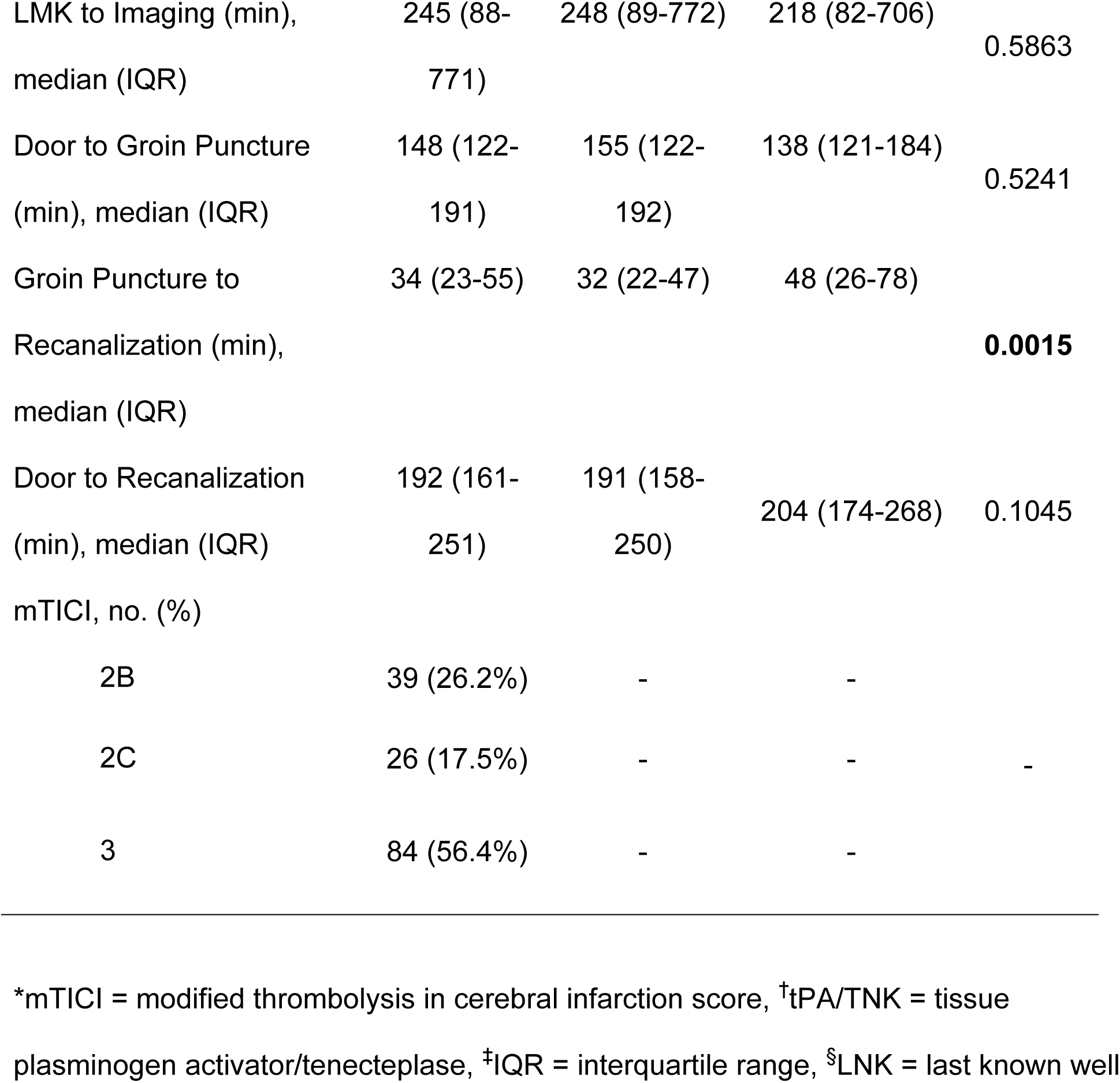
Comparison of interventions between patients recanalized to mTICI* 2B vs. 2C/3.

### Adjusted multivariate logistic regression analyses

After performing multivariate logistic regression analysis, the following parameters were identified as independent variables for prediction of excellent recanalization: lower admission NIHSS (aOR 0.93, p = 0.036), absence of history of DM (aOR 0.42, p = 0.050), and absence of prior stroke (aOR 0.27, p = 0.007), CBV index >= 0.7 (aOR 3.75, p = 0.007). Among interventional factors, aspiration-MT alone remained the only independent variables in predicting excellent recanalization (aOR 2.89, p = 0.012). See Table 4.

**4.**
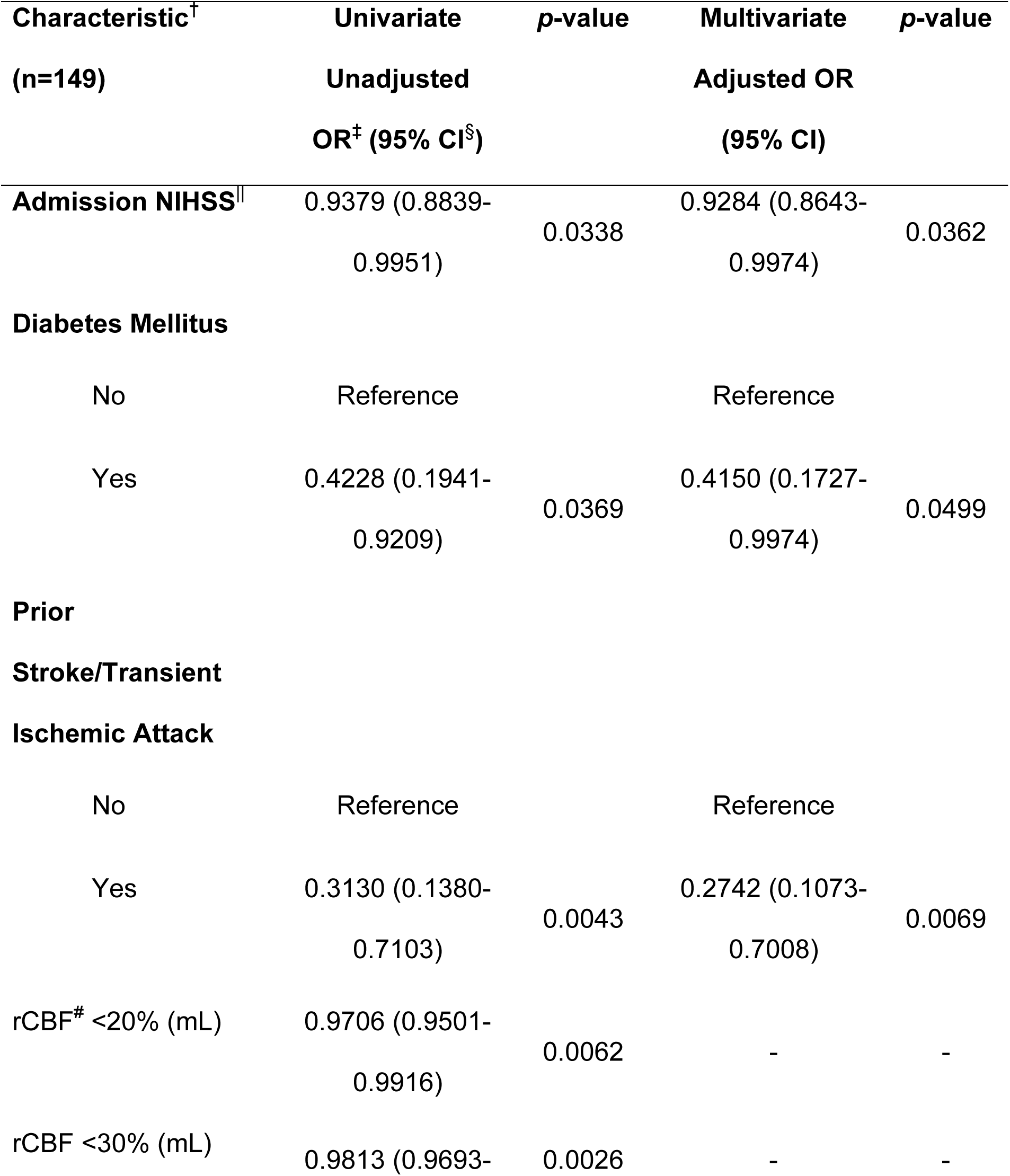
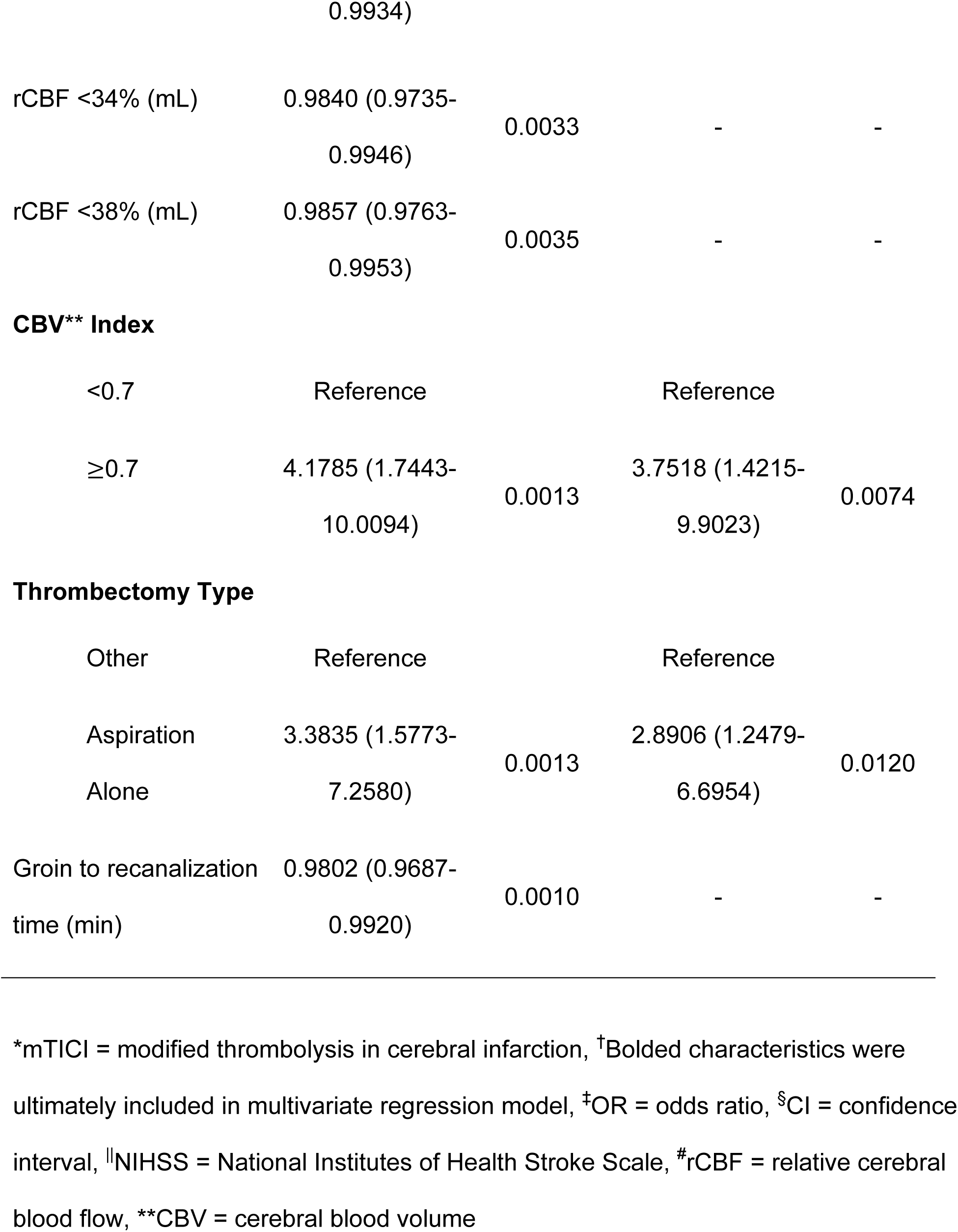
Significant unadjusted (univariate logistic regressions) and adjusted (stepwise multivariate logistic regression) odds ratio for outcome of mTICI* 2C/3 vs. 2B.

### ROC Analysis

ROC analyses showed that each of the variables ultimately included within the multivariate model had modest diagnostic performance in univariate logistic regression models for predicting excellent recanalization. These include (AUC, [95% CI)]: 0.62 [0.54 - 0.71] for rCBV index; 0.59 [0.51 - 0.68] for DM; 0.61 [0.53 - 0.70] for prior stroke; 0.62 [0.51 - 0.72] for admission NIHSS]; 0.65 [0.55 - 0.73] for aspiration-MT.

The multivariate logistic regression model composed of CBV Index >= 0.7, absence of DM, absence of prior stroke, admission NIHSS, and aspiration-MT demonstrated strong diagnostic performance with an AUC of 0.79 (95% CI: 0.68 - 0.86), p < 0.001) with a sensitivity of 0.94 (95% CI: 0.87-0.98) and specificity of 0.41 (95% CI: 0.01-0.60). See Figure 1.

**Figure 1:**
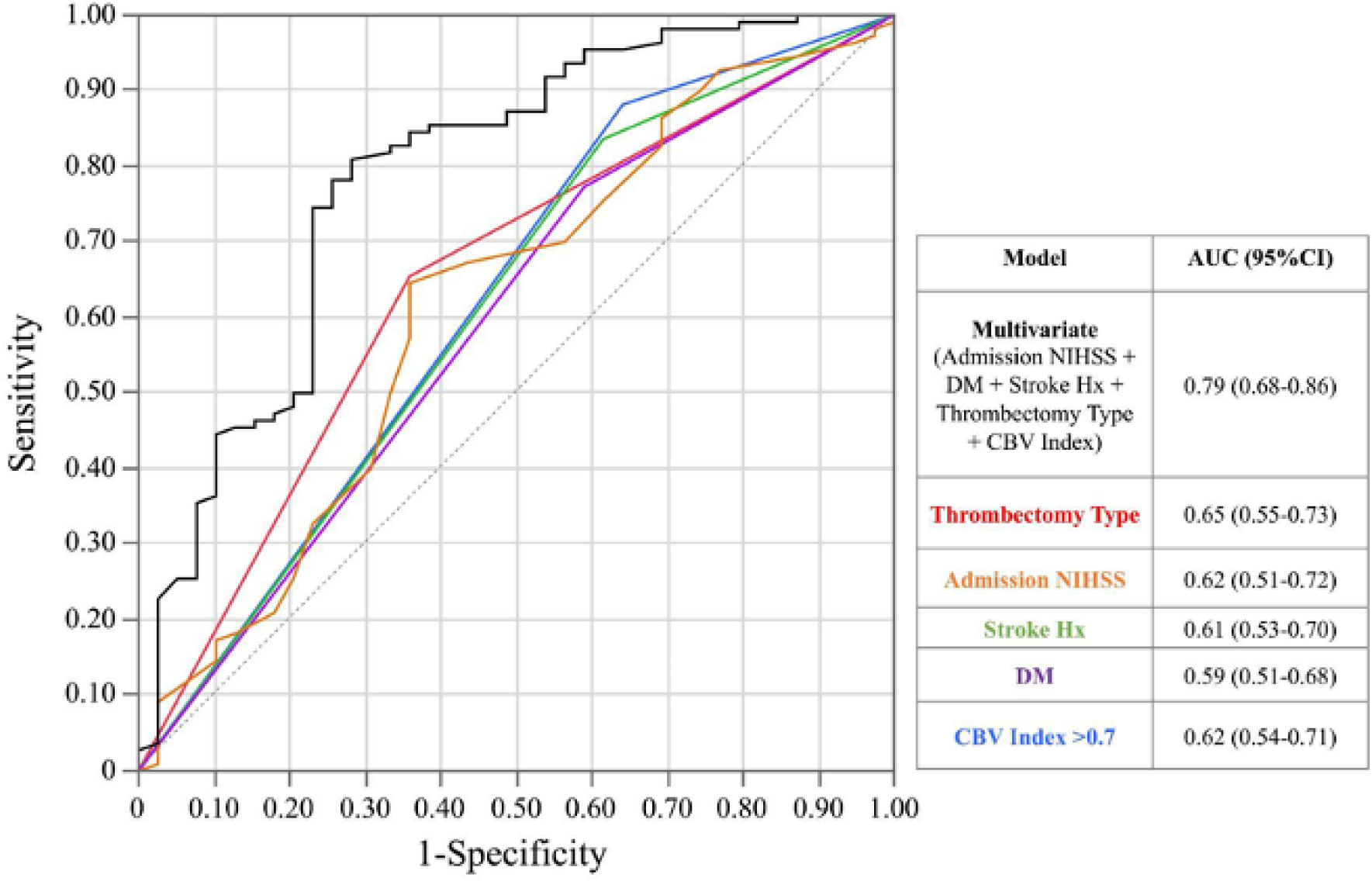
Receiver operator characteristics curve with area under the curve for dichotomized CBV index (>0.7), admission NIHSS, diabetes mellitus, prior stroke history, and aspiration thrombectomy as univariate factors and the combined multivariate model for outcome of mTICI 2C/3 versus 2B. Abbreviations: CBV = cerebral blood volume, NIHSS = National Institutes of Health Stroke Scale, mTICI = modified thrombolysis in cerebral infarction score, AUC = area under the curve, CI = confidence interval, DM = diabetes mellitus, Stroke Hx = stroke history.

## Discussion

Our study demonstrates that robust CS (as assessed by CBV index), aspiration alone, absence of DM and prior stroke, and lower initial stroke severity are all independent predictors of excellent recanalization amongst successfully recanalized AIS-LVO patients, with a strong combined predictive performance (AUC 0.79, p < 0.001). Our findings underscore the importance of pretreatment CS and treating these patients for their modifiable risk factors in order to maximize the likelihood of excellent recanalization, a strong indicator of favorable functional outcomes.^3–5^

A notable finding from our study is the utility of the CBV index not only as a reliable marker of CS but also as an independent predictor of excellent recanalization. The CBV index with a threshold of >0.7 has been previously validated as a predictor of CS^16, 17^, infarct growth rate, and clinical outcomes in AIS-LVO patients.^17–19^

In our study, we also identified that CBV index at an optimal threshold of >= 0.7 also predicts excellent recanalization, as a significantly higher percentage of CBV index >= 0.7 achieved excellent recanalization had CBV index >= 0.7 (88.2% versus 64% who achieved mTICI 2b, p<0.001). Moreover, a higher CBV index was strongly associated with achieving excellent recanalization (aOR 3.75, p = 0.007). The higher CBV index is a representation of collaterals as it is thought to indirectly represent blood volume that is preserved within the penumbra by compensatory responses such as autoregulation.

Our results also support prior work demonstrating the importance of CBV in maintaining robust CS and its subsequent effect of reperfusion. In the Interventional Management of Stroke III (IMS-III) trial, Liebeskind et al^20^ previously concluded that robust CS is associated with better recanalization and overall outcomes. In a subanalysis of the same trial, Vagal et al^21^ demonstrated an association with good CS and larger mismatch ratios on CT angiography/CTP evaluation, supporting the role of an autoregulatory response in maintaining tissue viability. Cortijo et al also reported that patients with a higher relative CBV have more robust CS and earlier recanalization.^22^ Most recently, Nael et al^23^ also highlighted the crucial role of CBV plays as a measure of collaterals. In this study, by incorporating CBV as an element of a multiparametric measure, described as the perfusion collateral index, they showed that higher baseline CS results in improved functional outcome and better reperfusion status following MT.^23^ It is thought that the pathophysiological mechanism behind increased CBV, as a component of robust CS, is due to prevention of stasis leading to additional distal occlusions from embolization.^24^ Furthermore, robust CS can also improve clearance of proximal thrombi^25^ and create pressure gradients across the occlusion that make thrombus retrieval more achievable.^24^

Besides CBV index, our results also revealed important findings about the interventional parameters as we showed that patients who achieved excellent recanalization had significantly fewer number of passes, were more likely to achieve recanalization through aspiration alone, and also had shorter times from groin puncture to recanalization (p = 0.002). These findings are undoubtedly intertwined as first pass effect^26^ leads to shorter time to recanalization^27, 28^, with both factors being associated with better outcomes. Direct aspiration is also increasingly linked to achieving excellent recanalization^11, 29–32^ compared to other techniques, and is likely also a contributing factor to the shorter puncture to recanalization times. However, in logistic regression analysis, aspiration-MT alone outperformed the other interventional measures and remained as the only contributing factor to our final model.

Our baseline clinical characteristics also revealed some notable findings. Unsurprisingly, patients with previous stroke, history of DM, and higher initial stroke severity were less likely to achieve excellent recanalization. Prior stroke is known to increase stroke recurrence^33^, and likely affects the robustness of CS. DM and hyperglycemia are also well researched predictors of poor outcomes in general for AIS-LVO patients.^34–36^ Additionally, higher stroke severity on admission is well established as a predictor of poorer outcomes.^37–39^

This study has limitations to acknowledge. It is limited by its retrospective design, which may lead to selection and unknown bias. Secondly, the mTICI score and interventional parameters were determined by the treating neurointerventionalist and were not validated by a central imaging core laboratory. This can lead to variability in reporting.^40^ Lastly, although CTP is readily available at comprehensive stroke centers, smaller rural sites and community hospitals may not have such capabilities.

These results have some important clinical implications. Our study further corroborates the importance of prevention of DM and stroke recurrence. Our results also emphasize the importance of robust CS and minimizing procedural time for optimizing likelihood of excellent recanalization. Larger scale prospective studies should be performed to ascertain the strength of these findings.

## Conclusion

CBV index>= 0.7, absence of DM and prior stroke, lower initial stroke severity and aspiration-MT are independent predictors of excellent recanalization in AIS-LVO patients.

## Data Availability

Data can be made available upon reasonable request.

## Acknowledgements

None

## Non-standard Abbreviations and Acronyms

| **Abbreviation** | **Definition** |
| --- | --- |
| AIS | Acute ischemic stroke |
| LVO | Large vessel occlusion |
| mTICI | Modified thrombolysis in cerebral infarction score |
| MT | Mechanical thrombectomy |
| CS | Collateral status |
| ICA | Internal carotid artery |
| M1 | M1 segment of middle cerebral artery |
| M2 | M2 segment of middle cerebral artery |
| CTP | CT perfusion |
| NIHSS | National Institutes of Health Stroke Scale |
| mRS | Modified Rankin score |
| DM | Diabetes mellitus |
| ASPECTS | Alberta stroke program early computed tomography score |
| Tmax | Time to maximum |
| rCBF | Relative cerebral blood flow |
| Hypoperfusion intensity ratio | HIR |
| CBV | Cerebral blood volume |
| ROC | Receiver operator curve |
| AUC | Area under the curve |

